# Unveiling the role of mosquito and human diel activity patterns in the risk of mosquito-borne disease infection

**DOI:** 10.1101/2024.05.29.24308139

**Authors:** Paulo C. Ventura, Andre B.B. Wilke, Jagadeesh Chitturi, Allisandra G. Kummer, Snigdha Agrawal, Chalmers Vasquez, Yaziri Gonzalez, Maria Litvinova, John-Paul Mutebi, Marco Ajelli

## Abstract

**Background:** Mosquito-borne pathogens are transmitted through bites of female mosquito vectors that are actively seeking hosts for a blood meal and hosts, when either of them is infectious. Different mosquito species have different preferences for the time of the day/night when they actively seek blood meals. In the United States, the encounters between mosquito vectors and human hosts primarily take place outdoors. Socioeconomic factors such as occupation and income are major determinants of the hour of the day and total amount of time spent outdoors by different population groups. The aim of this study is to quantify: i) diel variations in the level of human exposure to mosquito vectors, and ii) exposure heterogeneities by human population group.

**Methods:** We collected both diel activity data for two mosquito vector species (*Aedes aegypti* and *Culex quinquefasciatus*) and time-use data for the United States. Then, we analyzed the diel overlap between the two at the population level and by human population group.

**Results:** For both mosquito species, we found a substantial heterogeneity in their diel overlap with human outdoors activities. We estimated that the time periods with the highest risk of exposure to bites of *Ae. aegypti* are 7am-11am and 5pm-8pm, while the highest risk for *Cx. Quinquefasciatus* is 6am-7am and 6pm-9pm. Moreover, we found disparities in the exposure to mosquito vector species across different demographic groups. Workers with primarily outdoor occupations, males, and Hispanics/Latinos were shown to have higher levels of exposure as compared to the general population. In particular, we estimated that workers with primarily outdoor occupations were 7.50-fold (95%CI: 7.18-7.84) and 6.63-fold (95%CI: 6.09-7.35) more exposed to *Ae. aegypti* and *Cx. quinquefasciatus* than the general population, respectively.

**Conclusion:** This study serves as a steppingstone to quantify the risk of exposure to mosquito vector species in the United States. The obtained results can be instrumental for the design of public health interventions such as education campaigns, which could contribute to improve health and health equity.

## Introduction

With millions of cases and thousands of deaths reported every year, mosquito-borne diseases represent a major global public health threat [1,2]. Over the past fifty years, there has been a significant increase in global dengue incidence [3], with estimates pointing to 300 million infections every year [4]. The introduction of Zika in the Americas in 2015 resulted in an epidemic exceeding 1 million reported cases [5,6]. Malaria remains a substantial public health burden, causing over 500,000 deaths annually [7]. Albeit on a smaller scale, the United States has witnessed a surge in local outbreaks of mosquito-borne diseases in recent years [8,9]. For instance, Miami-Dade County, Florida, has experienced local dengue outbreaks for the last three consecutive years (i.e., 2022-2024) [9]. For the first time in the last 20 years, the Centers for Disease Control and Prevention (CDC) reported locally acquired cases of *Plasmodium vivax* malaria in Florida and Texas in 2023 [10]. Moreover, between 1999 and 2022, the United States reported 56,575 cases of West Nile virus infection, resulting in 25,777 hospitalizations and 2,776 fatalities, with the largest outbreak ever reported taking place in Maricopa County, Arizona, in 2021. [11].

Mosquito-borne pathogens are transmitted through bites of female mosquito vectors that are actively seeking hosts for a blood meal and hosts, when either of them is infectious [12]. Different mosquito species have different preferences for the time of the day/night when they actively seek blood meals. For example, *Aedes aegypti* (a primary vector for dengue, Zika, and chikungunya viruses) is a crepuscular species [13,14]; on the other hand, *Culex quinquefasciatus* (a primary vector for West Nile virus and St. Louis Encephalitis Virus) is mostly active at night [13,14]. In the United States, the encounters between mosquito vectors and human hosts primarily occur outdoors [10,14–16]. Socioeconomic factors such as occupation and income are major determinants of the hour of the day and total amount of time spent outdoors leading to differences between population groups (e.g., gender, racial/ethnical) [17,18].

The aim of this study is to quantify: i) diel variations in the level of human exposure to mosquito vectors, and ii) exposure heterogeneities by human population group. To this aim, we collected both diel activity data for two mosquito vector species (*Ae. aegypti* and *Cx. quinquefasciatus*) and (human) time-use data for the United Stated. These data are used to quantify the diel overlap between the two mosquito vector species and the human population both at the population level and by human population group. By shedding light on the overlap between human and mosquito vector activities, our study could be instrumental for the design of public health interventions such as education campaigns, which could contribute to improve health and health equity.

## Methods

### Mosquito diel activity data

Diel activity data for *Ae. aegypti* and *Cx. quinquefasciatus* was retrieved from our previous studies [13,14]. Briefly, in those studies, mosquitoes were collected by BG-Sentinel 2 traps (Biogents AG, Regensburg, Germany) baited with BG Lures and dry ice. Mosquitoes were collected once a month every hour for 96 consecutive hours in four locations within two study sites: Miami-Dade County, Florida, and Brownsville, Texas. Collected mosquitoes were kept under refrigeration until identified to species on chill tables according to entomological keys [19]. For details on the data collection, we refer the reader to the two original studies [13,14].

### Human diel activity data

To infer human outdoor diel activity patterns, we used data from the American Time Use Survey (ATUS) [20]. The ATUS data is collected from a sample of the US resident population that is representative by race/ethnicity, gender, and occupation. Each participant reports their activities according to a list of 17 first-tier categories, which is further subdivided into two additional tier categories. For each participant, the data is collected for a 24-hour period with minute resolution. The participants can optionally inform the location type of the activity from a list of 24 single-tier categories. This data is openly available along with detailed socio-demographic information for each participant [20]. We collected time-used data for every participant between 2014 and 2019. For each collected year of data, we restricted our analysis to the period between May and September, as it corresponds to the period when the majority of locally acquired infections of mosquito-borne diseases in the contiguous United States take place [9,11].

### Estimation of the time spent outdoors

Following the methodology of Hoehne et al. [21], we classified their activities as taking place indoors, outdoors, or at an unknown/mixed location. When the participant reported the location where the activity took place, we coded the time spent in that location as either indoors or outdoors depending on the location type. Instead, when a respondent reported to be working, the activity was classified based on the respondent’s occupation (see Supplementary Material for details).

### Estimation of the exposure to mosquito vectors

To estimate the risk of exposure to bites of mosquito vectors by hour of the day relative to 6pm (which is used as reference hour), we used the following equation:

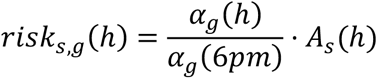

where:

- *s* represents the analyzed mosquito species, namely *Aedes aegypti* or *Culex quinquefasciatus*.
- *g* represents the human population group, namely the general population, occupation (outdoor; indoor/mixed), race/ethnicity (Hispanic; Non-Hispanic White; Non-Hispanic Black; Non-Hispanic Other), gender (female; male), or income (low; middle; high).
- *h* represents the hour of the day.
- 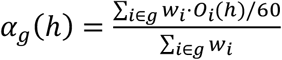 is the probability that an individual of group *g* is outdoor at hour of the day *h*. In the equation for *α*_*g*_(*h*), *i* represents a survey participant that belongs to population group *g*; *w*_*i*_ represents the weight of participant *i* provided by the ATUS survey to obtain a US-representative population; *O*_*i*_(*h*) represents the number of minutes that participant *i* spent outdoors during hour *h*.
- *A* 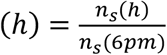 is the proportion of female mosquitoes of species *s* that are actively seek hosts for blood meal at hour *h* relative to 6pm. In the equation for *A*_*s*_(*h*), *n*_*s*_(*h*) represents the number of collected mosquitoes of species *s* at hour *h*.

Note that as reference hour, we used 6pm since at that time both the focus mosquito species show some level of host seeking behavior and human hosts show some level of outdoor activity.

To estimate the risk of exposure to bites of mosquito vectors by human population group relative to the general population, we used the following equation:

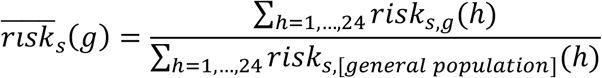

### Estimation of the uncertainty

To estimate uncertainty in mosquito diel activity, we model the collection of specimens as a binomial process. Each specimen in the entire mosquito population can be trapped and collected with a certain probability. Assuming that the actual number of specimens in the population is much larger than the number of collected ones (i.e., that the probability of collection is small), we can approximate this to a Poisson process. Under this assumption, the uncertainty for each number of collected specimens was estimated as the 95% IQR of a Poisson distribution whose average is the value observed in the field.

To estimate uncertainty on human diel activity, we performed a bootstrap sampling. In each iteration, we resampled the set of individuals and performed the classification analysis with the new ensemble.

The uncertainty on the estimates presented in this study are the result of 1,000 samples from a Poisson distribution and 1,000 independent bootstrap iterations.

## Results

### Mosquito and human activity patterns by hour of the day

Both analyzed mosquito species show well-defined species-specific diel activity patterns (Fig. 1A, 1B). Females of *Ae. aegypti* are highly active in seeking hosts for blood meals between 7am and 10am, and between 6pm and 9pm (Fig. 1A). We found the hour of maximum activity to be 8 pm, when we estimated a proportion of active female mosquitoes of 2.49 (95%CI: 2.39-2.57) as compared to 6pm; the hour of lowest activity was 12 am, when the proportion of active female mosquitoes as compared to 6pm was of 0.21 (95%CI: 0.17-0.24).

**Figure 1.**
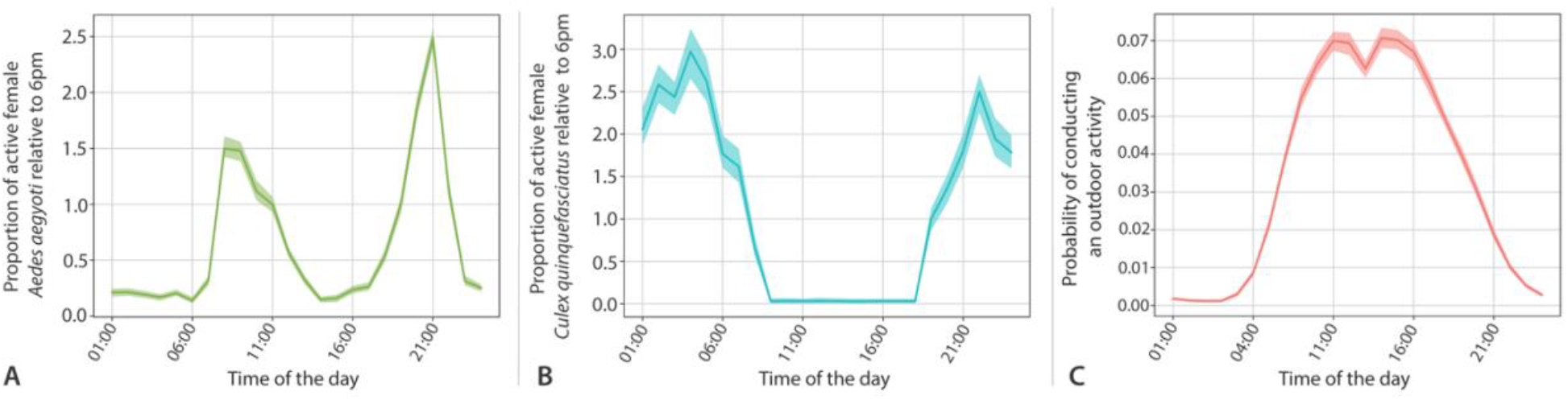
Mosquito and human diel activity patterns. **A** Proportion of female *Ae. aegypti* that are actively seek hosts for blood meal at a given hour of the day relative to 6pm. **B** Same as A but for *Cx. quinquefasciatus*. **C** Probability that a US resident is conducting an outdoor activity at a given hour of the day.

Females of *Cx. quinquefasciatus* were primarily active in seeking hosts for blood meals between 6pm and 7am (Fig. 1B). The hour of maximum activity was 3 am, with an estimated proportion of active female mosquitoes of 2.97 (95%CI: 2.65-3.24) as compared to 6pm; no substantial activity was estimated between 8 am and 5pm.

We found that human activity patterns follow a diurnal cycle, with daytime hours marked by heightened outdoor activity levels due to work, education, and social engagements (Fig. 1C). We found the period with highest outdoor activity level to span from 9 am to 4 pm when 6-7% of the population spends time outdoors.

### Risk of exposure to mosquito vectors by hour of the day

By studying the overlap between human and mosquito activities, we found a well-defined pattern with two peaks per day of high level of exposure (Fig. 2A-B). However, these peaks take place at different time for the two analyzed mosquito species. Specifically, we found high exposure levels to *Ae. aegypti* between 7am-11am and 5pm-8pm (Fig. 2A), with maximum exposure taking place at 8 am (exposure risk: 2.05, 95%CI: 1.91-2.20, relative to 6 pm). For *Cx. quinquefasciatus* (Fig. 2B), the periods of highest exposure found between 6am-7am and 6pm-9pm, with maximum exposure taking place at 7 pm (exposure risk: 1.02, 95%CI: 0.86-1.16, relative to 6 pm).

**Figure 2.**
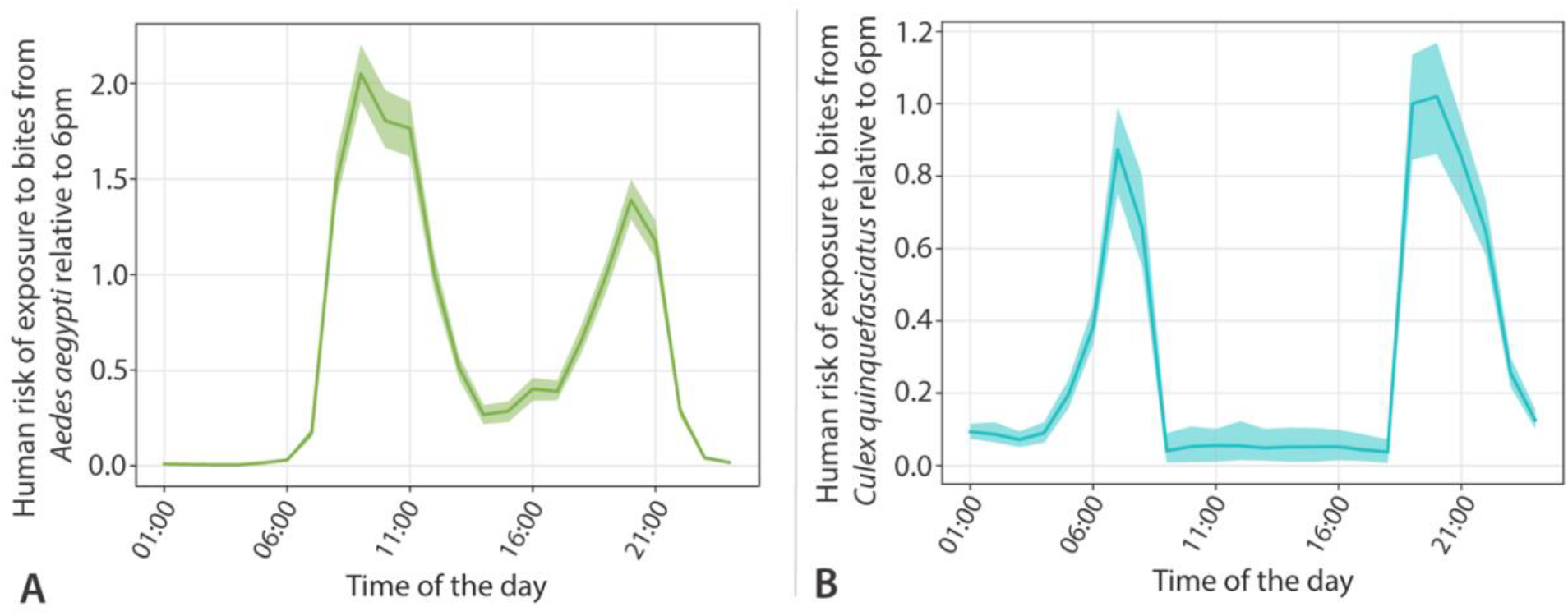
Human exposure to mosquito vector species by hour of the day. **A** Human risk of exposure to bites of *Ae. aegypti* by hour of the day relative to 6pm. **B** Same as A but for *Cx. quinquefasciatus*.

### Human population heterogeneities in the amount of time spent outdoors

The amount of outdoor activity of the US population is highly heterogeneous, with 75% of the population spending less than 1 hour per day outdoors, while 22% spending at least one hour and 2% spending more than 8 hours (Fig. 3A). This high heterogeneity is associated with socio-economic characteristics of the population (Fig. 3B). Occupation type was the largest source of heterogeneity, where primarily outdoors workers such as people employed in the landscaping and construction sectors spent a substantially larger amount of time outdoors as compared to other occupation types: 6.23 hours per day (95%CI: 5.97-6.44) vs. 0.62 hours per day (95%CI: 0.61-0.63). By dividing the US population in different racial/ethnical groups, we found that Hispanics/Latinos spent the highest amount of time outdoors (1.07 hours per day, 95%CI: 0.98-1.15), followed by Non-Hispanic (NH) Whites (0.85 hours per day, 95%CI: 0.83-0.87), NH others (0.57 hours per day, 95%CI: 0.50-0.63), and NH Blacks (0.46 hours per day, 95%CI: 0.42-0.50). When considering gender, we estimated that males spent more time outdoors (1.20 hours per day, 95%CI: 1.16-1.24) than females (0.47 hours per day, 95%CI: 0.45-0.48), possibly associated with difference in occupation. Finally, we did not find substantial differences when dividing the population by income level.

**Figure 3.**
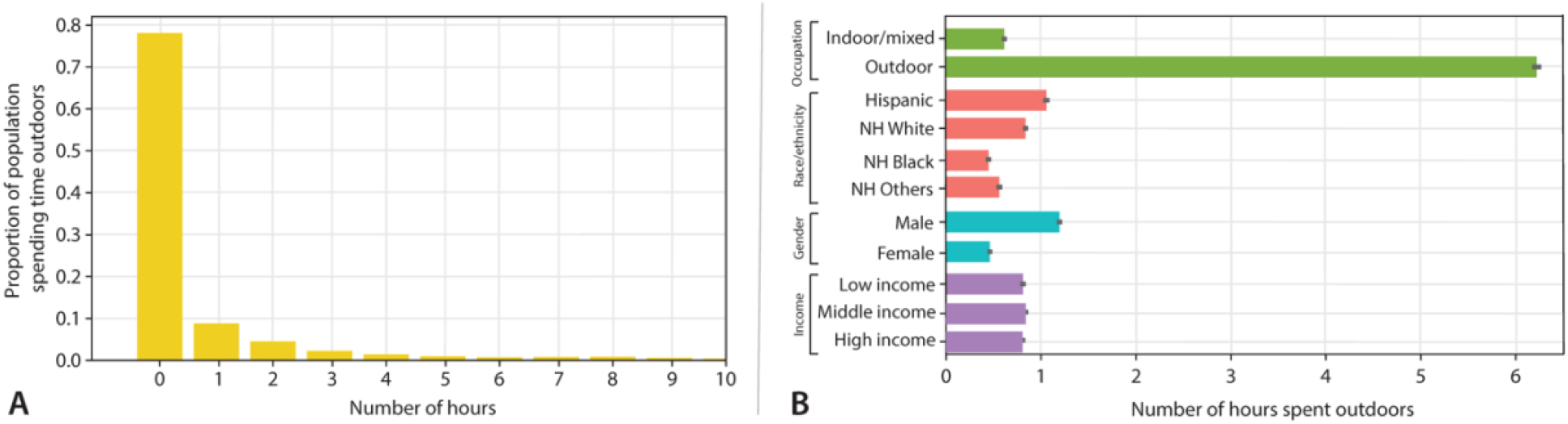
Heterogeneity in the amount of time spent outdoors by the US population. **A** Distribution of the amount of time spent outdoors per day by the US population. **B** Mean number of hours spent outdoors per day for different population groups. Outdoor occupations include: Farming, fishing, forestry, construction and extraction occupations. Respondents whose family income was under the 33% quantile of their state were classified as low income, while those over the 67% quantile were classified as high income, and all others were classified as middle income.

### Human population heterogeneities in the risk of exposure to mosquito vectors

The highest levels of exposure to mosquito vector species were found to be associated with the type of occupation. Workers with primarily outdoor occupations yielded the highest levels of exposure being 7.50 times (95%CI: 7.18-7.84) more exposed to *Ae. aegypti* and 6.63 times (95%CI: 6.09-7.35) more exposed to *Cx. quinquefasciatus* in comparison to the general population. Occupations that are primarily indoors have a substantially lower risk of exposure to both mosquito species (Fig. 4). The level of human exposure to mosquito vectors varied greatly by race/ethnicity. Hispanics had the highest exposure to mosquitoes being 1.32 times (95%CI: 1.22-1.44) more exposed to *Ae. aegypti* and 1.36 times (95%CI: 1.17-1.56) more exposed to *Cx. quinquefasciatus* in comparison to the general population. NH Black bore the lowest exposure to mosquitoes to both mosquito species as compared to the general population: 0.55 (95%CI: 0.50-0.61) for *Ae. aegypti* and 0.63 (95%CI: 0.56-0.71) for *Cx. quinquefasciatus* (Fig. 4). Males are estimated to be 1.45 (95%CI: 1.40-1.50) times more exposed to *Ae. aegypti* and 1.39 times (95%CI: 1.30-1.48) more exposed to *Cx. quinquefasciatus* in comparison to the general population, whereas we found a reduced exposure for females (0.58, 95%CI: 0.56-0.60 for *Ae. aegypti* and 0.64, 95%CI 0.60-0.69 for *Cx. quinquefasciatus*) (Fig 4). Finally, no significant difference in the risk of exposure to both analyzed mosquito vectors species was found between income level groups (Fig. 4). While the risk of exposure to the two analyzed vector species is highly heterogenous with the US population, the time of the day at which different population groups have higher exposure to mosquito vectors is consistent between human population group (see Supplementary Material, Figs. S1 and S2).

**Figure 4.**
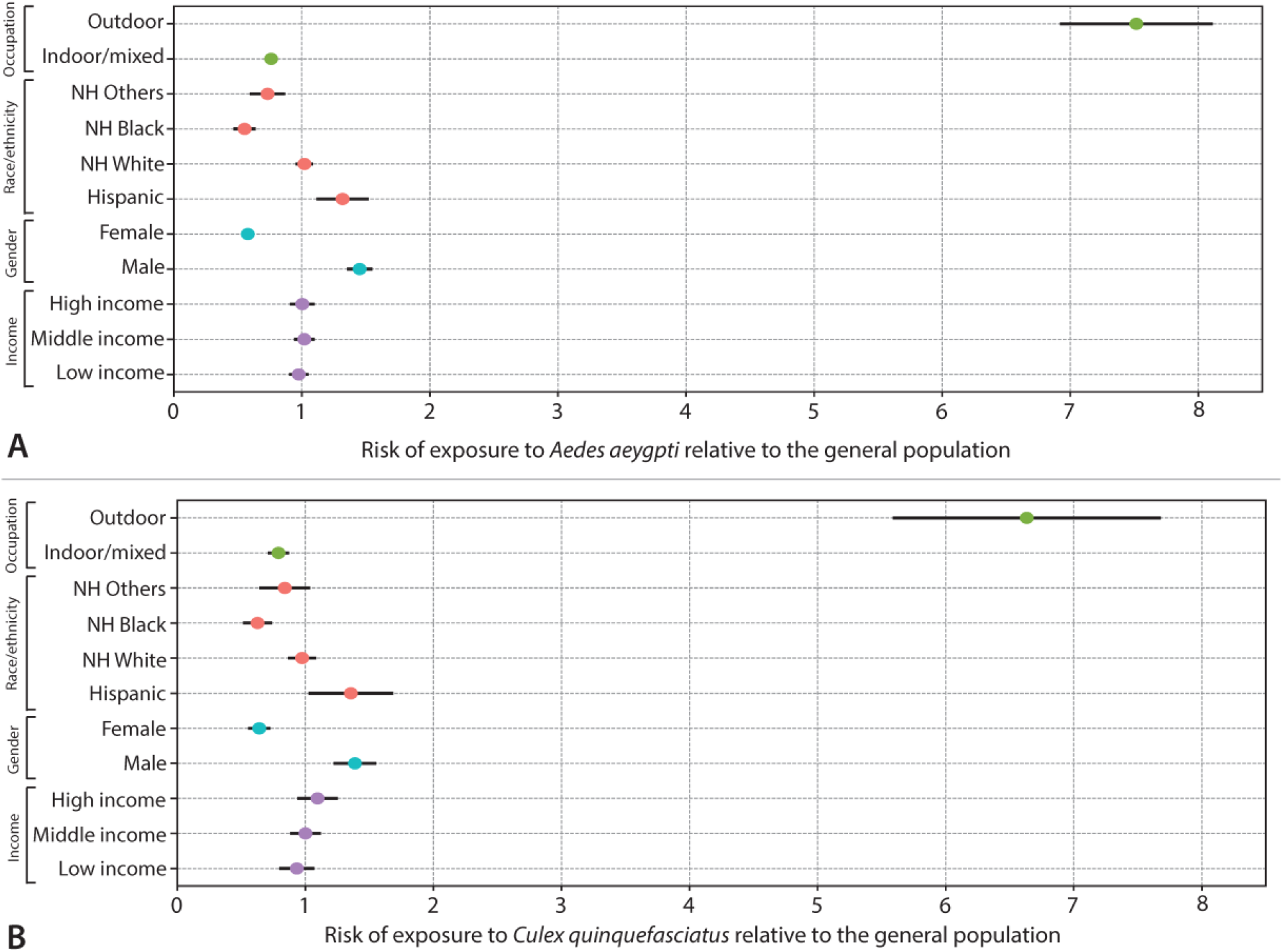
Human population heterogeneities in the risk of exposure to mosquito vectors. **A** Risk of exposure to *Ae. aegypti* by human population group relative to the general population. **B** As A but for *Cx. quinquefasciatus*.

## Discussion

Our results show substantial heterogeneity in the diel activity patterns of both mosquitoes and humans, encompassing periods of low activity as well as peaks of heightened activity. Furthermore, mosquito and human diel activity peaks overlap at specific hours during the day shedding light on when the risk of being exposed to mosquito vector species is higher. The heterogeneity in the amount of time spent outdoors by different human population groups is associated with substantial disparities in the exposure to mosquito vector species across different demographic groups. We found that workers with primarily outdoor occupations, Hispanics/Latinos, and males have higher levels of exposure to mosquito vector species. This nuanced understanding underscores the complexity of human exposure to mosquitoes and the risk of mosquito-borne infection.

Mitigating outbreaks of mosquito-borne diseases is resource-intensive and entails social and environmental costs [22,23]. Thus, determining the influence of diel activity patterns on the risk of mosquito-borne infection and the effectiveness of public health interventions are essential to plan resource allocation to maximize the effectiveness of outbreak preparedness and response and minimize their side effects [24]. For example, implementing adulticide spraying interventions aimed at controlling *Ae. aegypti* populations during peak activity periods can yield entomological outcomes that are fivefold more effective compared to conducting the same intervention during periods of low activity [13]. This highlights that mosquito diel activity patterns have a strong impact on the entomological outcome of adulticide spraying. Additionally, the likelihood and magnitude of outbreak occurrences were shown to substantially diminish by when interventions align with peak mosquito activity periods [25]. The findings presented in this study can further our understanding of the effectiveness of alternative public health interventions including educational campaigns on the hours of the day with highest exposure to mosquito vector species. Moreover, our findings can be instrumental to identify the audience of educational campaigns to maximize their effectiveness.

In this study, we analyzed the daily activity of mosquitoes from May to September when humans are most active outdoors in the US, at the national level, exposed to mosquito bites. This allowed us to gain insights into their behavioral patterns during periods of peak activity [9,11]. We acknowledge that different areas may have distinct climate and environmental cues that can affect human and mosquito diel activity patterns differently. Moreover, mosquito and human diel activity patterns often fluctuate throughout the year following shifts in sunrise and sunset times. Future studies should focus on furthering our understanding of how behavioral traits change through the year and by geographical areas. Another limitation of our study is outdoor activities were inferred from time use data, which was not purposedly designed for this objective. For this reason, some of the activities were classified as taking place in a mixed or unknown environment. Future studies should address this limitation and extensive sensitivity analyses showing the impact of different time-use classification are warranted.

In conclusion, this study serves as a steppingstone to quantify the risk of exposure to mosquito-borne pathogens and their transmission dynamics. Ultimately, this framework based on the analysis of the overlap between human and mosquito activities has the potential to inform the development of targeted public health interventions, ultimately improving health and health equity.

## Data Availability

All data produced are available online at
https://www.bls.gov/tus/ and doi: 10.1371/journal.pntd.0011074.

## Declarations

### Ethical Approval

The Institutional Review Board at Indiana University has considered this research to be exempt from review (IRB #23194).

