## Supplementary_Material for "Unveiling the role of mosquito and human diel activity patterns in the risk of mosquito-borne disease infection"

### Methods Details

#### Classification of the time use data

To quantify human diel activity in terms of indoor and outdoor presence, we used data from the American Time Use Survey (ATUS) and Current Population Survey (CPS), both being performed by the USA Bureau of Labor Statistics (BLS). The CPS is a survey of households in the United States that collects a wide range of information about employment, unemployment, hours of work, earnings, and people not in the labor force. The ATUS is a continuous survey that measures how individuals in the United States allocate their time across various activities. For the ATUS, participants are randomly selected from households that have completed their eighth month of interviews for the CPS. Each respondent is interviewed once, detailing their activities from the previous day, including where they were and whom they were with. The respondents are asked to report the start and end times of each activity with minute precision during a contiguous period of 24 hours. The CPS and ATUS surveys also collect demographic information such as sex, race, age, educational attainment, occupation, income, marital status, and the presence of children in the household. The anonymized microdata is publicly available at https://www.bls.gov/tus/.

From the CPS and ATUS microdata, we estimate patterns of outdoor presence by classifying the individual records following a detailed criterion. Specifically, we use the following data:

- **Activity type**, as reported by the respondents following a three-tier multiple choice classification system (TRCODE field). The system has 17 first-tier categories, subdivided into hundreds of third-tier categories.
- **Activity location type**, which is optionally reported for each activity using a single-tier multiple choice system with 24 valid options plus 2 options for unspecified places (TEWHERE field).
- **Respondent's primary and secondary jobs** in the day of the diary, to classify "at work" activities. We used the primary (TRDTOCC1 field) and secondary (TRDTOCC2 field) occupation types, which follow the two-tier Census Bureau's Occupation Classification System. For this purpose, we used the first-tier code only.

We first created classification tables for each of the above data. For the activity types, we used the criterion from Hoehne et al. [1]. For the activity location types, we constructed a table by discerning locations that are most likely indoors or outdoors, setting as "unknown" locations that can be mixed or cannot be clearly specified (see table S1). Similarly, for the occupation types, we classified those which are most likely to be predominantly performed indoors or outdoors, setting as "unknown" those that are mixed or cannot be clearly specified (see table S2).

Using these tables, we classified each record of an activity performed by a respondent. The procedure for each record is described as follows:

- **Initialization**: The record is first set as "Unclassified" (not to be confused with "Unknown").
- **Activity type**: If the activity type code (TRCODE) is found in the classification table, the record is attributed to the corresponding class – "Indoor", "Outdoor" or "Unknown".
- **Activity location type**: If the activity location code (TEWHERE) is found in the corresponding classification table *and* it is either "Indoor" or "Outdoor", the record is set to the corresponding class, overriding the previous classification.
- **Work activities**: If the record indicates that the participant is at work (TRCODE == 050101 for primary job and TRCODE == 050102 for secondary job), then the record is set according to the job classification – "Indoor", "Outdoor" or "Unknown". This does not override the previous steps, since "at work" codes are not in the classification table.
- **Unclassified**: If the record still holds the "Unclassified" label, it is set as "Unknown".

Once every activity record of an individual's diary was classified as "Indoor", "Outdoor" or "Unknown", we expanded the records into a minute-resolution time series of diel activity. This specifies, at each minute of the 24-hour period, if the individual was indoor, outdoor or at an unknown-type location.

To construct the results presented in this work, the individual data was aggregated using the representativity weights provided in the ATUS data (TUFINLWGT field). For example, to determine the intensity of outdoor presence for all residents of the United States at a given time, we added the weights of all individuals that were outdoors at that time and divided by the sum of weights of all individuals. We also combined the ATUS datasets from year 2014 to 2019, resulting in 6 years of data in total for a bigger sample size. The choice of years avoids mixing versions of the survey with significant methodological differences, while also avoiding biases and issues due to the COVID-19 pandemic. To combine data from different years, we renormalized the individual weights such that the ensemble of each year contributed equally to the overall aggregation.

Finally, to estimate the confidence intervals on the human diel activity patterns, we performed a bootstrap analysis by resampling the entire set of individuals. For the results in the main text, we performed 1000 independent resampling runs.

**Table S1.** Classification of the location categories as indoor, outdoor and unknown.

| ATUS Location Code (TEWHERE) | ATUS Location Category | Indoor/Outdoor Classification |
|---|---|---|
| 1 | Respondent's home or yard | Unknown |
| 2 | Respondent's workplace | Unknown |
| 3 | Someone else's home | Unknown |
| 4 | Restaurant or bar | Unknown |
| 5 | Place of worship | Unknown |
| 6 | Grocery store | Indoor |
| 7 | Other store/mall | Unknown |
| 8 | School | Unknown |
| 9 | Outdoors away from home | Unknown |
| 10 | Library | Unknown |
| 11 | Other place | Unknown |
| 12 | Car, truck, or motorcycle (driver) | Indoor |
| 13 | Car, truck, or motorcycle (passenger) | Indoor |
| 14 | Walking | Outdoor |
| 15 | Bus | Indoor |
| 16 | Subway/train | Indoor |
| 17 | Bicycle | Outdoor |
| 18 | Boat/ferry | Unknown |
| 19 | Taxi/limousine service | Indoor |
| 20 | Airplane | Indoor |
| 21 | Other mode of transportation | Unknown |
| 30 | Bank | Indoor |
| 31 | Gym/health club | Unknown |
| 32 | Post Office | Unknown |
| 89 | Unspecified place | Unknown |
| 99 | Unspecified mode of transportation | Unknown |

**Table S2**. Classification of occupation codes, using the 2010 Census Occupation Classification system, as indoor, outdoor and unknown.

| Detailed Occupation Code | 2010 Occupation Description | Indoor/Outdoor Classification |
|---|---|---|
| 1 | Management Occupations | Indoor |
| 2 | Business and financial operations occupations | Indoor |
| 3 | Computer and mathematical science occupations | Indoor |
| 4 | Architecture and engineering occupations | Unknown |
| 5 | Life, Physical, and social science occupations | Indoor |
| 6 | Community and social service occupations | Unknown |
| 7 | Legal occupations | Indoor |
| 8 | Education, training, and library occupations | Indoor |
| 9 | Arts, design, entertainment, sports, and media occupations | Unknown |
| 10 | Healthcare practitioner and technical occupations | Indoor |
| 11 | Healthcare support occupations | Indoor |
| 12 | Protective service occupations | Unknown |
| 13 | Food preparation and serving related occupations | Indoor |
| 14 | Building and grounds cleaning and maintenance occupations | Unknown |
| 15 | Personal care and service occupations | Indoor |
| 16 | Sales and related occupations | Indoor |
| 17 | Office and administrative support occupations | Indoor |
| 18 | Farming, fishing, and forestry occupations | Outdoor |
| 19 | Construction and extraction occupations | Outdoor |
| 20 | Installation, maintenance, and repair occupations | Unknown |
| 21 | Production occupations | Unknown |
| 22 | Transportation and material moving occupations | Unknown |

### Additional Results

#### Exposure by hour of the day for different population groups

We show how the risk of exposure to mosquito vectors evolves during the hours of the day and for different population groups. For both mosquito species, the two-peak pattern is consistent between population groups, showing that optimal hours of exposure are the same among the population (Fig. S1 and S2).

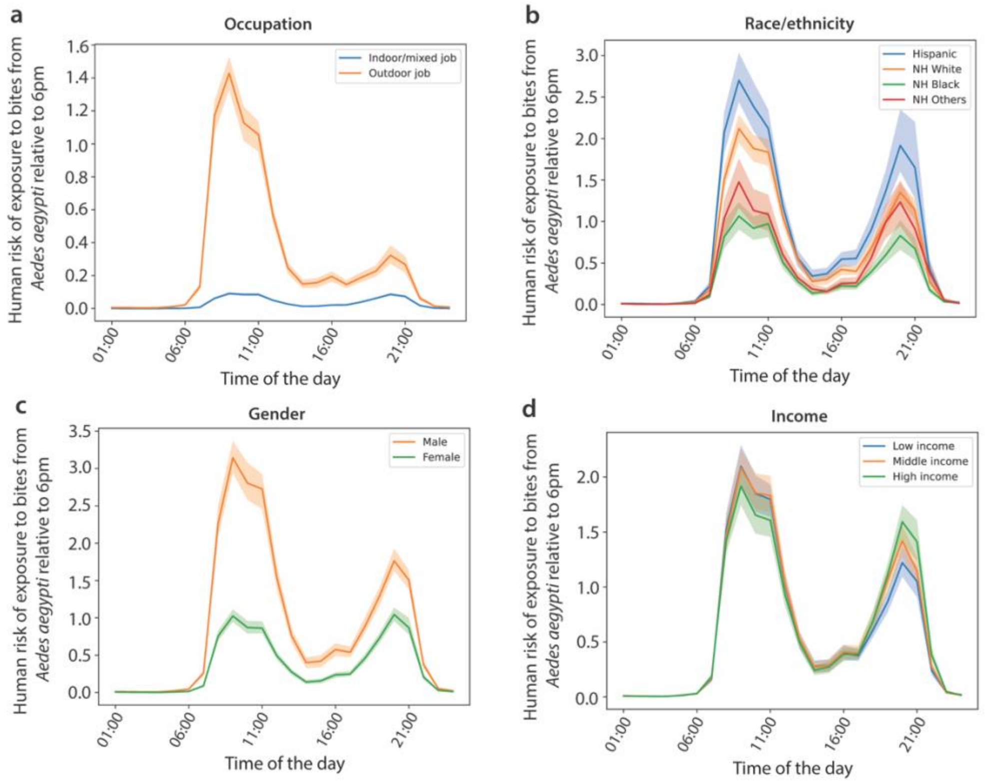


**Figure S1**. **Human exposure to *Ae. aegypti* by hour of the day and human population group. A** Human exposure to *Ae. aegypti* by hour of the day relative to 6pm by occupation. **B** As A but for race/ethnicity. **C** As A but for gender. **D** As A but for income.

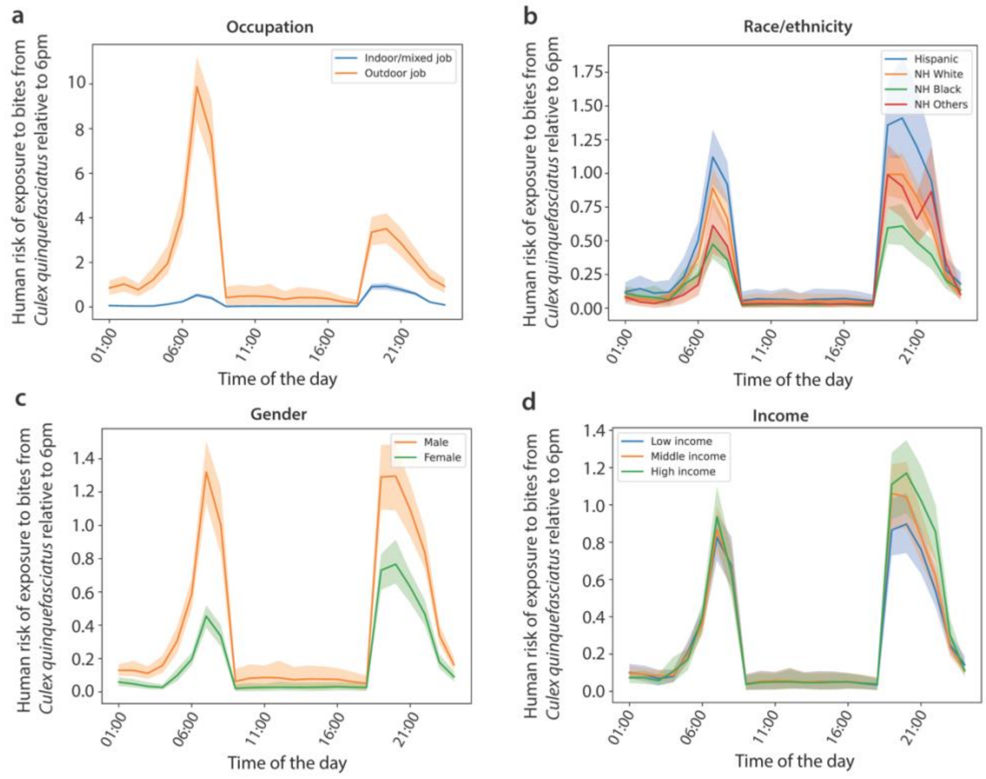


**Figure S2**. **Human exposure to *Cx. quinquefasciatus* by hour of the day and human population group. A** Human exposure to *Cx. quinquefasciatus* by hour of the day relative to 6pm by occupation. **B** As A but for race/ethnicity. **C** As A but for gender. **D** As A but for income.
